# Transplantation of solid organs recovered from deceased donors recently infected by SARS-CoV-2 in the United States

**DOI:** 10.1101/2022.06.05.22276008

**Authors:** Jonathan M. Czeresnia, Helen Tsai, Maria Ajaimy, Clara Y. Tow, Snehal R. Patel, Ulrich P. Jorde, Shivank Madan, Vagish Hemmige

**Author notes:** Correspondence to: Vagish Hemmige, M.D., M.S., 3411 Wayne Avenue, Suite 4E, Bronx, NY, United States of America, 10467.

## Abstract

The COVID-19 pandemic has reduced access to solid organ transplantation, compounding organ shortages and waitlist mortality. A continued area of uncertainty is the safety of transplanting organs recovered from SARS-CoV-2 infected donors, as autopsies of patients who died with COVID-19 show that the virus can be found in extra-pulmonary organs^1^. Case reports and series on transplantation of these organs have been published ^2, 3^, but population-level data is lacking.

We queried a national transplant database for recipients of organs recovered from donors recently infected by SARS-CoV-2. For organs with more than 50 cases, these were then propensity-score matched at a ratio of 1:10 to similar recipients of organs recovered from donors who tested negative for SARS-CoV-2 (controls). Data were extracted from the Scientific Registry of Transplant Recipients (SRTR - v2203 - updated March 2022), which collects detailed information on all solid organ transplants in the United States since 1986.

Cases were defined as adult (≥ 18 years) recipients of organs recovered from deceased donors who tested positive for SARS-CoV-2 by nasopharyngeal or lower respiratory sample polymerase chain reaction or antigen assay within 7 days of organ transplantation. Multiple organ transplants were excluded.

There were 775 kidney, 330 liver, 123 heart, 44 kidney-pancreas, 16 lung, 5 pancreas, and 3 small bowel transplants of organs recovered from 393 deceased donors recently infected by COVID-19. For kidney, liver, and heart transplants, Kaplan-Meier curves of both overall and graft survival at 90 days were similar between cases and controls.

Our data shows that transplanting kidneys, livers, and hearts recovered from deceased donors recently infected by SARS-CoV-2 was not associated with increased recipient mortality or worse graft-survival. This should help transplant providers make decisions regarding acceptance of these organs, and counsel transplant candidates on the safety of receiving them. The limited number of kidney-pancreas, lung, pancreas, and intestinal cases precludes significant conclusions for these organs. Our data also strongly supports the notion that donors with recent COVID-19 infection should not be automatically excluded from the donor pool. The limited number of kidney-pancreas, lung, pancreas, and intestinal cases precludes significant conclusions for these organs.

Limitations include lack of data on donor infection timeline and estimates of viral load (PCR cycle thresholds), description of donor COVID-19 symptomatology at organ procurement, donor or recipient vaccination or prior COVID-19 infection status, which are not tracked in the database. We did not have information regarding transmission of COVID-19 to transplant recipients. Future analysis of updated versions of the database should help address. Our data strongly support the notion that donors with recent COVID infection should not be automatically excluded from the donor pool. Prospective studies are needed to confirm our findings and provide insights on optimal post-transplant management of these recipients.

## 1. Introduction

The COVID-19 pandemic has drastically affected the field of solid organ transplantation (SOT), far beyond the direct effects of SARS-CoV-2 itself on transplant recipients. The unprecedented strain on healthcare systems initially caused a sharp decline in SOTs^4, 5^. After transplant programs had time to adjust to our new reality, there was a rebound in the number of SOTs, with some programs even exceeding pre-pandemic levels ^6^. A continued area of uncertainty is the safety of transplanting organs recovered from SARS-CoV-2 infected donors. Autopsy studies of patients who died with or of COVID-19 have found evidence of viral invasion of most, if not all, extra-pulmonary tissues^1^, raising concerns that SARS-CoV-2 may be transmitted with organ transplantation.

Case reports and series of these transplants have been published for several organs, showing mostly good outcomes and no evidence of transplant transmission of COVID-19^7, 8, 9, 10, 11^, but population-level data is lacking.

## 2. Methods

To address this issue, we queried the Scientific Registry of Transplant Recipients (SRTR - v2203 - last reported transplants: 3/2/2022) for adult (≥ 18 years) recipients of organs recovered from deceased donors who tested positive for SARS-CoV-2 by nasopharyngeal or lower respiratory polymerase chain reaction (PCR) or antigen assay within 7 days of organ transplantation. RStudio v1.4 running base R v4.1.2 (Bird Hippie) with previously described packages *(eg. matchit, tableone, survival, survminer)* attached was used for matching and statistical analysis. In the interest of scientific transparency and reproducibility, source code is provided at the end of this article.

This study used data from the SRTR. The SRTR data system includes data on all donor, wait-listed candidates, and transplant recipients in the US, submitted by the members of the Organ Procurement and Transplantation Network (OPTN). The Health Resources and Services Administration (HRSA), U.S. Department of Health and Human Services provides oversight to the activities of the OPTN and SRTR contractors.

### a. Case selection

SRTR does not include data on severity, onset, or viral kinetics of SARS-CoV-2 infection. Donor cause of death is grouped into categories which do not allow for conclusions regarding if a donor died from COVID-19 or with COVID-19, though it is safe to assume that donors with severe infection would be declined. Most tests reported to the database were nasopharyngeal or lower respiratory PCRs (50,277 of 53,079) followed by antigen assays (973 of 53,079). Serologies and tests reported as “other” were excluded. As most reported specimens were obtained close to enlistment and not onset of disease, we decided to consider donors who tested positive for COVID-19 by PCR or antigen tests up to 7 days prior to transplantation as being recipients of “COVID-19 positive organs”. Other time cutoffs, such as increasing the range of time between testing positive and transplantation, or limiting time between procurement and positive test, did not significantly alter outcomes. We then found adult (≥ 18y) recipients of these kidneys, livers, and hearts, which we defined as cases. There were fewer than 50 recipients of lung, kidney-pancreas, pancreas, and intestinal transplants, which are briefly described below but not included in our statistical analysis. There were no cases of heart-lung transplants.

### b. Matching

Matched demographic characteristics were the same across organs. Use of indication for transplant, severity scoring systems, and waitlist status varied depending on which data were consistently available and pertinent for that specific organ. All controls underwent transplantation between 2020 and 2022 and were above 18 years of age at time of transplant. Multi-organ transplants were excluded. Matched characteristics are as follows:

- All organs: donor age, recipient age at transplant, recipient race/ethnicity, recipient gender.

In addition:

- Kidneys: Kidney Donor Profile Index (KDPI), indication for transplant.
- Livers: all cases were matched for donor heart-beating status. Status 1A patients, those with diagnoses related to HCC, and patients with a diagnosis of acute liver failure were matched independently. For all others, Model for End-Stage Liver Disease-Na (MELD-Na) score was used.
- Hearts: waitlist status at time of transplant, donor gender.

Propensity score matching was performed approximately for continuous variables and exactly for categorical ones at a ratio of 1:10. Repeated controls were not allowed. Standardized mean differences between cases and controls was below 10% for matched characteristics, indicating adequate balance. Comparative characteristics of cases and controls can be found in Table 1.

**Table 1:**
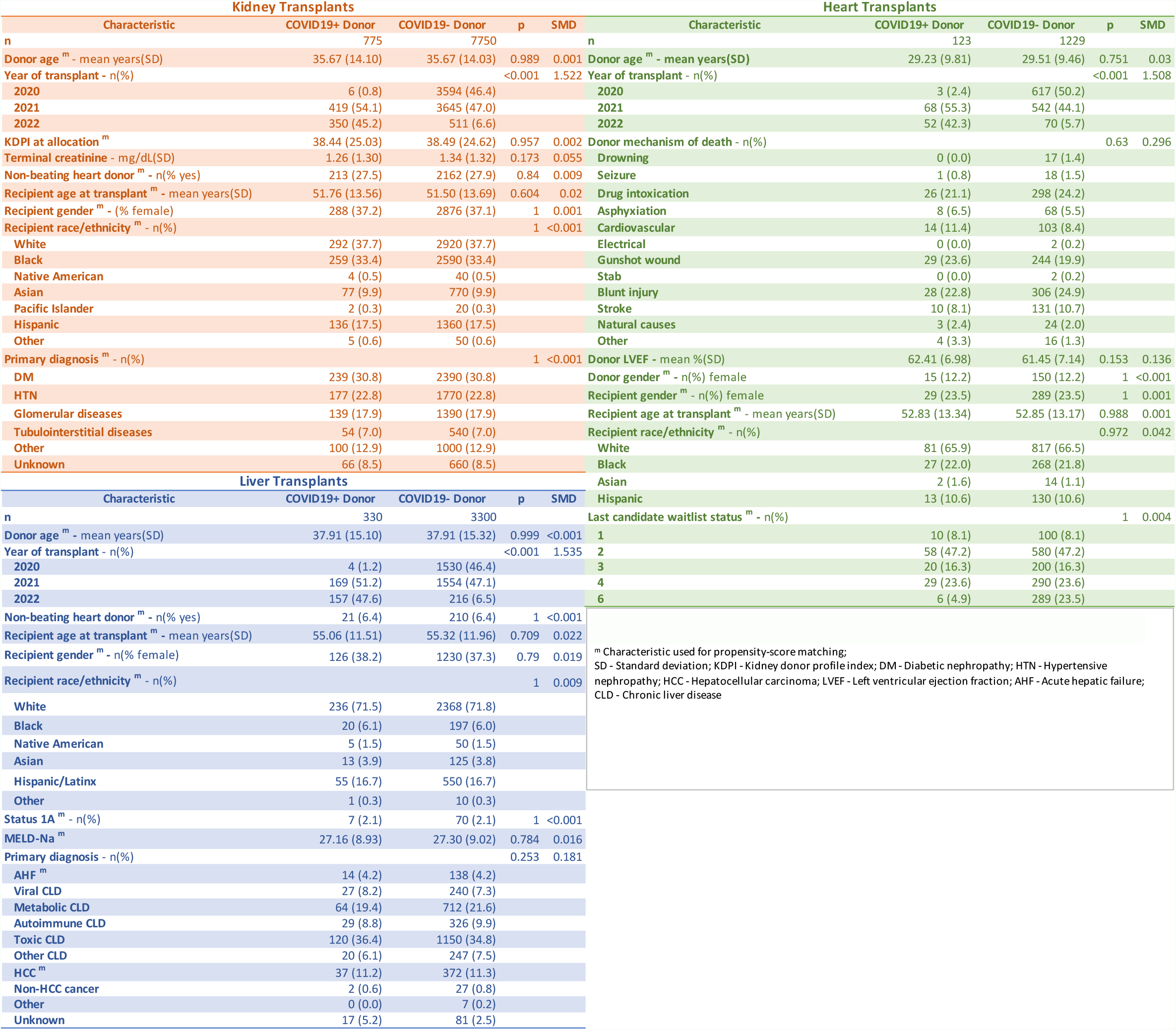
Characteristics of kidney, liver, and heart transplants of recipients of organs recovered from donors recently infected by SARS-CoV-2, compared to recipients of organs of non-infected donors.

Kidney Transplants
| Characteristic | COVID19+ Donor | COVID19- Donor | p | SMD |
| --- | --- | --- | --- | --- |
| n | 775 | 7750 |  |  |
| Donor age <sup>m</sup> - mean years(SD) | 35.67 (14.10) | 35.67 (14.03) | 0.989 | 0.001 |
| Year of transplant - n(%) |  |  | <0.001 | 1.522 |
| 2020 | 6 (0.8) | 3594 (46.4) |  |  |
| 2021 | 419 (54.1) | 3645 (47.0) |  |  |
| 2022 | 350 (45.2) | 511 (6.6) |  |  |
| KDPI at allocation <sup>m</sup> | 38.44 (25.03) | 38.49 (24.62) | 0.957 | 0.002 |
| Terminal creatinine - mg/dL(SD) | 1.26 (1.30) | 1.34 (1.32) | 0.173 | 0.055 |
| Non-beating heart donor <sup>m</sup> - n(% yes) | 213 (27.5) | 2162 (27.9) | 0.84 | 0.009 |
| Recipient age at transplant <sup>m</sup> - mean years(SD) | 51.76 (13.56) | 51.50 (13.69) | 0.604 | 0.02 |
| Recipient gender <sup>m</sup> - (% female) | 288 (37.2) | 2876 (37.1) | 1 | 0.001 |
| Recipient race/ethnicity <sup>m</sup> - n(%) |  |  | 1 | <0.001 |
| White | 292 (37.7) | 2920 (37.7) |  |  |
| Black | 259 (33.4) | 2590 (33.4) |  |  |
| Native American | 4 (0.5) | 40 (0.5) |  |  |
| Asian | 77 (9.9) | 770 (9.9) |  |  |
| Pacific Islander | 2 (0.3) | 20 (0.3) |  |  |
| Hispanic | 136 (17.5) | 1360 (17.5) |  |  |
| Other | 5 (0.6) | 50 (0.6) |  |  |
| Primary diagnosis <sup>m</sup> - n(%) |  |  | 1 | <0.001 |
| DM | 239 (30.8) | 2390 (30.8) |  |  |
| HTN | 177 (22.8) | 1770 (22.8) |  |  |
| Glomerular diseases | 139 (17.9) | 1390 (17.9) |  |  |
| Tubulointerstitial diseases | 54 (7.0) | 540 (7.0) |  |  |
| Other | 100 (12.9) | 1000 (12.9) |  |  |
| Unknown | 66 (8.5) | 660 (8.5) |  |  |

Liver Transplants
| Characteristic | COVID19+ Donor | COVID19- Donor | p | SMD |
| --- | --- | --- | --- | --- |
| n | 330 | 3300 |  |  |
| Donor age <sup>m</sup> - mean years(SD) | 37.91 (15.10) | 37.91 (15.32) | 0.999 | <0.001 |
| Year of transplant - n(%) |  |  | <0.001 | 1.535 |
| 2020 | 4 (1.2) | 1530 (46.4) |  |  |
| 2021 | 169 (51.2) | 1554 (47.1) |  |  |
| 2022 | 157 (47.6) | 216 (6.5) |  |  |
| Non-beating heart donor <sup>m</sup> - n(% yes) | 21 (6.4) | 210 (6.4) | 1 | <0.001 |
| Recipient age at transplant <sup>m</sup> - mean years(SD) | 55.06 (11.51) | 55.32 (11.96) | 0.709 | 0.022 |
| Recipient gender <sup>m</sup> - n(% female) | 126 (38.2) | 1230 (37.3) | 0.79 | 0.019 |
| Recipient race/ethnicity <sup>m</sup> - n(%) |  |  | 1 | 0.009 |
| White | 236 (71.5) | 2368 (71.8) |  |  |
| Black | 20 (6.1) | 197 (6.0) |  |  |
| Native American | 5 (1.5) | 50 (1.5) |  |  |
| Asian | 13 (3.9) | 125 (3.8) |  |  |
| Hispanic/Latinx | 55 (16.7) | 550 (16.7) |  |  |
| Other | 1 (0.3) | 10 (0.3) |  |  |
| Status 1A <sup>m</sup> - n(%) | 7 (2.1) | 70 (2.1) | 1 | <0.001 |
| MELD-Na <sup>m</sup> | 27.16 (8.93) | 27.30 (9.02) | 0.784 | 0.016 |
| Primary diagnosis - n(%) |  |  | 0.253 | 0.181 |
| AHF <sup>m</sup> | 14 (4.2) | 138 (4.2) |  |  |
| Viral CLD | 27 (8.2) | 240 (7.3) |  |  |
| Metabolic CLD | 64 (19.4) | 712 (21.6) |  |  |
| Autoimmune CLD | 29 (8.8) | 326 (9.9) |  |  |
| Toxic CLD | 120 (36.4) | 1150 (34.8) |  |  |
| Other CLD | 20 (6.1) | 247 (7.5) |  |  |
| HCC <sup>m</sup> | 37 (11.2) | 372 (11.3) |  |  |
| Non-HCC cancer | 2 (0.6) | 27 (0.8) |  |  |
| Other | 0 (0.0) | 7 (0.2) |  |  |
| Unknown | 17 (5.2) | 81 (2.5) |  |  |

| Characteristic | COVID19+ Donor | COVID19- Donor | p | SMD |
| --- | --- | --- | --- | --- |
| n | 123 | 1229 |  |  |
| Donor age <sup>m</sup> - mean years(SD) | 29.23 (9.81) | 29.51 (9.46) | 0.751 | 0.03 |
| Year of transplant - n(%) |  |  | <0.001 | 1.508 |
| 2020 | 3 (2.4) | 617 (50.2) |  |  |
| 2021 | 68 (55.3) | 542 (44.1) |  |  |
| 2022 | 52 (42.3) | 70 (5.7) |  |  |
| Donor mechanism of death - n(%) |  |  | 0.63 | 0.296 |
| Drowning | 0 (0.0) | 17 (1.4) |  |  |
| Seizure | 1 (0.8) | 18 (1.5) |  |  |
| Drug intoxication | 26 (21.1) | 298 (24.2) |  |  |
| Asphyxiation | 8 (6.5) | 68 (5.5) |  |  |
| Cardiovascular | 14 (11.4) | 103 (8.4) |  |  |
| Electrical | 0 (0.0) | 2 (0.2) |  |  |
| Gunshot wound | 29 (23.6) | 244 (19.9) |  |  |
| Stab | 0 (0.0) | 2 (0.2) |  |  |
| Blunt injury | 28 (22.8) | 306 (24.9) |  |  |
| Stroke | 10 (8.1) | 131 (10.7) |  |  |
| Natural causes | 3 (2.4) | 24 (2.0) |  |  |
| Other | 4 (3.3) | 16 (1.3) |  |  |
| Donor LVEF - mean %(SD) | 62.41 (6.98) | 61.45 (7.14) | 0.153 | 0.136 |
| Donor gender <sup>m</sup> - n(%) female | 15 (12.2) | 150 (12.2) | 1 | <0.001 |
| Recipient gender <sup>m</sup> - n(%) female | 29 (23.5) | 289 (23.5) | 1 | 0.001 |
| Recipient age at transplant <sup>m</sup> - mean years(SD) | 52.83 (13.34) | 52.85 (13.17) | 0.988 | 0.001 |
| Recipient race/ethnicity <sup>m</sup> - n(%) |  |  | 0.972 | 0.042 |
| White | 81 (65.9) | 817 (66.5) |  |  |
| Black | 27 (22.0) | 268 (21.8) |  |  |
| Asian | 2 (1.6) | 14 (1.1) |  |  |
| Hispanic | 13 (10.6) | 130 (10.6) |  |  |
| Last candidate waitlist status <sup>m</sup> - n(%) |  |  | 1 | 0.004 |
| 1 | 10 (8.1) | 100 (8.1) |  |  |
| 2 | 58 (47.2) | 580 (47.2) |  |  |
| 3 | 20 (16.3) | 200 (16.3) |  |  |
| 4 | 29 (23.6) | 290 (23.6) |  |  |
| 6 | 6 (4.9) | 289 (23.5) |  |  |

<sup>m</sup> Characteristic used for propensity-score matching; SD - Standard deviation; KDPI - Kidney donor profile index; DM - Diabetic nephropathy; HTN - Hypertensive nephropathy; HCC - Hepatocellular carcinoma; LVEF - Left ventricular ejection fraction; AHF - Acute hepatic failure; CLD - Chronic liver disease

### c. Outcomes

Overall survival and graft loss at 90 days were chosen as primary outcomes because these are reliably tracked in the database. True incidence of transplant COVID-19 transmission could not be accurately assessed, as detailed courses of each case is not available. The most specific data we found is presented below.

Curves comparing overall survival and incidence of graft loss (calculated as time to first event) between cases and controls were generated using the Kaplan-Meier (KM) method (Figure 1). Likelihood ratios were calculated by univariate Cox regression (Table 2). For secondary outcomes expected to be closely tracked (e.g. occurrence of rejection during transplant admission, delayed graft function), missing values were interpreted as negative, unless follow up time was equal to zero (only transplant event itself recorded), in which case patients were excluded. Other missing data were excluded.

**Table 2:**
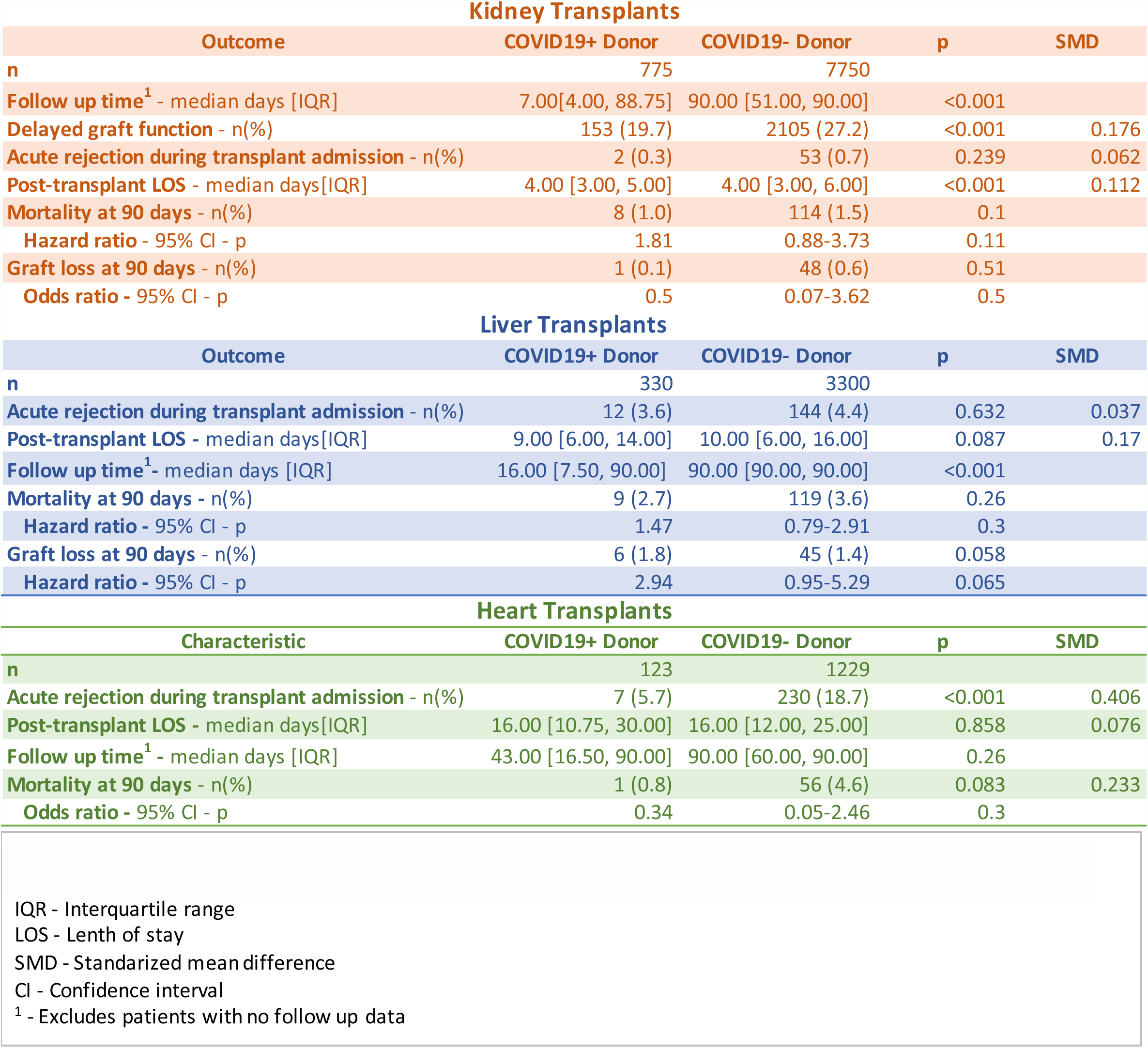
Outcomes of kidney, liver, and heart transplantation in recipients of organs recovered from donors recently infected by SARS-CoV-2, compared to recipients of organs of non-infected donors.

Kidney Transplants
| Outcome | COVID19+ Donor | COVID19- Donor | p | SMD |
| --- | --- | --- | --- | --- |
| n | 775 | 7750 |  |  |
| Follow up time<sup>1</sup> - median days [IQR] | 7.00[4.00, 88.75] | 90.00 [51.00, 90.00] | <0.001 |  |
| Delayed graft function - n(%) | 153 (19.7) | 2105 (27.2) | <0.001 | 0.176 |
| Acute rejection during transplant admission - n(%) | 2 (0.3) | 53 (0.7) | 0.239 | 0.062 |
| Post-transplant LOS - median days[IQR] | 4.00 [3.00, 5.00] | 4.00 [3.00, 6.00] | <0.001 | 0.112 |
| Mortality at 90 days - n(%) | 8 (1.0) | 114 (1.5) | 0.1 |  |
| Hazard ratio - 95% CI - p | 1.81 | 0.88-3.73 | 0.11 |  |
| Graft loss at 90 days - n(%) | 1 (0.1) | 48 (0.6) | 0.51 |  |
| Odds ratio - 95% CI - p | 0.5 | 0.07-3.62 | 0.5 |  |

Liver Transplants
| Outcome | COVID19+ Donor | COVID19- Donor | p | SMD |
| --- | --- | --- | --- | --- |
| n | 330 | 3300 |  |  |
| Acute rejection during transplant admission - n(%) | 12 (3.6) | 144 (4.4) | 0.632 | 0.037 |
| Post-transplant LOS - median days[IQR] | 9.00 [6.00, 14.00] | 10.00 [6.00, 16.00] | 0.087 | 0.17 |
| Follow up time<sup>1</sup> - median days [IQR] | 16.00 [7.50, 90.00] | 90.00 [90.00, 90.00] | <0.001 |  |
| Mortality at 90 days - n(%) | 9 (2.7) | 119 (3.6) | 0.26 |  |
| Hazard ratio - 95% CI - p | 1.47 | 0.79-2.91 | 0.3 |  |
| Graft loss at 90 days - n(%) | 6 (1.8) | 45 (1.4) | 0.058 |  |
| Hazard ratio - 95% CI - p | 2.94 | 0.95-5.29 | 0.065 |  |

| Characteristic | COVID19+ Donor | COVID19- Donor | p | SMD |
| --- | --- | --- | --- | --- |
| n | 123 | 1229 |  |  |
| Acute rejection during transplant admission - n(%) | 7 (5.7) | 230 (18.7) | <0.001 | 0.406 |
| Post-transplant LOS - median days[IQR] | 16.00 [10.75, 30.00] | 16.00 [12.00, 25.00] | 0.858 | 0.076 |
| Follow up time<sup>1</sup> - median days [IQR] | 43.00 [16.50, 90.00] | 90.00 [60.00, 90.00] | 0.26 |  |
| Mortality at 90 days - n(%) | 1 (0.8) | 56 (4.6) | 0.083 | 0.233 |
| Odds ratio - 95% CI - p | 0.34 | 0.05-2.46 | 0.3 |  |

IQR - Interquartile range
LOS - Length of stay
SMD - Standardized mean difference
CI - Confidence interval
<sup>1</sup> - Excludes patients with no follow up data

**Figure 1:**
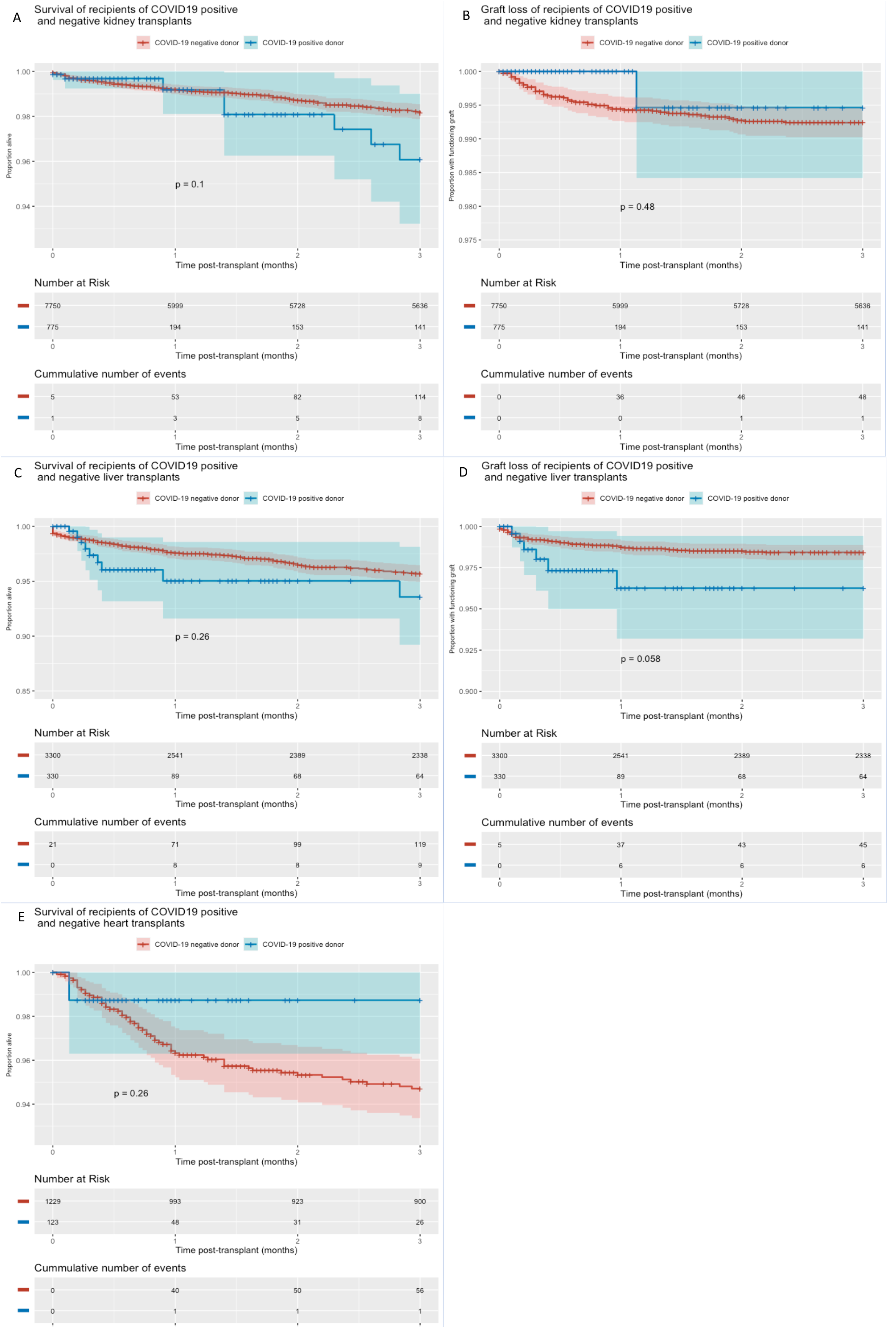
Kaplan-Meier curves of recipients of transplants involving organs recovered from donors recently infected by SARS-CoV-2 and of recipients of transplants from donors who tested negative for COVID-19. p calculated by log rank test. Curve of graft loss in hearts omitted, as there were no occurrences of graft loss that did not cause death in the recipients of COVID19+ organs. Shaded areas in plots correspond to 95% confidence intervals.

### d. Follow up time

The dataset included a significant number of very recent transplants in the group that received COVID-19 positive organs. These either only had entries of the transplant event itself, with no follow up data, or only of the transplant admission. Cases with no follow up data were excluded when calculating median follow up times. As the statistical methods we used account for differences in follow up time, patients without follow up had no weight in calculating correlations. The number of patients at risk and cumulative number of events at each point are provided below the curves (Figure 1). Matching for follow up time was not pursued, as it would have resulted in selection bias.

## 2. Results

There were 775 kidney, 330 liver, 123 heart, 44 kidney-pancreas, 16 lung, 5 pancreas, and 3 small bowel transplants of organs recovered from 393 deceased donors recently infected by SARS-CoV-2.

## a. Kidneys

There were 775 kidney transplants from recently infected SARS-CoV-2 donors identified. 213 of these had no follow up data recorded. The number of these transplants increased between 2020 and 2022 (6 performed in 2020, 419 performed in 2021, and 350 performed in the first three months of 2022). The terminal creatinine for cases and controls was not significantly different. The leading recipient primary diagnosis for transplant was diabetes mellitus.

Table 1 summarizes outcomes of 775 kidney transplant recipients who received allografts from SARS-CoV-2 positive donors and 7750 kidney transplant recipients who received allografts from SARS-CoV-2 negative donors. Delayed graft function, defined as the need of dialysis within the first week after transplant, was lower in cases than in controls (19.7% vs. 27.2%, p<0.001).

There were no significant differences in rates of rejection episodes occurring during the index admission (0.3% vs. 0.7%, p=0.24). Compared to matched controls, kidney recipients of SARS-CoV-2 positive donors had similar 90-day mortality (1% vs. 1.5%, p=0.11, HR 1.81 [95% CI 0.88 – 3.73]) (Figure 1a). Moreover, recipient graft loss within 90 days of transplant were not significantly different between the two groups (0.3% vs. 1.1%, p=0.5, HR 0.50 [95% CI 0.07 – 3.62]) (Figure 1a). The median follow-up period was lower for cases than controls (7 vs. 90 days p<0.001), though 141 cases who were followed for 90 days. Among patients who received positive organs, there were 8 deaths, only one of which had COVID-19 listed as cause. As this occurred 42 days post-transplant, and the patient was discharged from the hospital in the interim, COVID-19 was presumably acquired in the community. There were 2 occurrences of graft loss among cases, one due to primary non-function and the second of unknown cause.

### b. Livers

We identified 330 cadaveric livers with recent COVID-19 infection that were transplanted between 2020-2022 (Table 1). 95 of these had no follow up data reported. The number of liver transplants from SARS-CoV-2 positive donors increased over time, as 4 (1.1%) of solitary liver transplants were performed in 2020 compared to 169 (51.2%) in 2021 and 157 (47.6%) over the course of 3 months in 2022 (time that SRTR data was extracted). Seventy-eight percent of solitary liver recipients were transplanted for chronic liver conditions, with toxic liver disease comprising that largest percentage of patients (36.4% cases and 34.8 controls). Rare cases of acute liver failure meeting criteria for status 1A listing were observed (2.1% cases and controls), and approximately 10% of cases and controls were transplanted for hepatocellular carcinoma (HCC) via standard HCC exception points. The average MELD-Na at the time of transplant for both cases and controls was approximately 27.

The most meaningful outcomes, 90-day mortality and graft loss, were similar between recipients of organs from SARS-CoV-2 positive or negative donors. Compared to matched controls, liver recipients of SARS-CoV-2 positive donors had similar 90-day mortality (2.7% vs. 4.0%, p=0.26, HR 1.41 [95% CI 0.74-2.91]) (Figure 1c). Ninety-day graft loss trended towards statistically higher in COVID+ recipients (1.8 vs. 1.4, p=0.058, HR 2.24 [95% CI 0.95-5.29]), but was not clinically significant (Figure 1d). There was also no difference in episodes of acute cellular rejection during the index admission (4.9% vs. 4%, p=0.581). The median follow up time for cases was significantly lower than that for controls (9 days versus 90 days, p<0.001), with 64 cases who were followed for 90 days.

Among patients who received positive organs, there were 9 deaths, none of which had COVID-19 listed as a cause. There were 6 cases of graft loss, two of which had “infection” listed as causes. More granular data was not available.

### c. Hearts

There were 123 heart transplants from recently infected SARS-CoV-2 donors, 44 of which had no follow up data reported, and 26 who were followed for 90 days. The frequency of these transplants increased significantly since the onset of the COVID-19 pandemic (3 in 2020 vs. 68 in 2021 vs. 52 in early 2022). Most donors died of gunshot wounds (23.6%), followed by blunt injury (22.8), and drug intoxication (21.1%). Mean donor left ventricular ejection fraction was 62.4% (SD 6.98). Most donors (88.8%) and recipients (76.5%) were men. Most recipients were waitlist status 2 prior to transplant (47.2%).

Incidence of rejection during index admission was lower in the recipients of COVID-19 positive hearts (5.7% vs 18.7%, p<0.001). Length of stay for index admission was similar. There was one death among patients who received positive organs, caused by primary graft failure. There were no other occurrences of graft loss. Mortality at 90 days was similar between cases and matched controls (Figure 1E).

### d. Other organs

- Lungs: 16 patients met the definition of cases. Median follow up time was 41.5 days (IQR 8.25-71.25). There were no deaths reported;
- Pancreas: 5 patients, followed for a median of 50 days (IQR 41-90). There were no deaths reported;
- Bowel: 3 patients, followed for a median of 50 days (IQR 45.5-90). There were no deaths reported;
- Kidney-pancreas: 44 patients met the definition of cases. Median follow up time was 9.5 days (IQR 2.25 - 47.77). There were no deaths reported.
- Heart-lung: there were no cases.

## 6. Discussion

Our data shows that transplanting kidneys, livers, and hearts recovered from deceased donors recently infected by SARS-CoV-2 was not associated with increased recipient mortality or worse graft-survival (Figure 1, Table 2). This should help transplant providers make decisions regarding acceptance of these organs, and counsel transplant candidates on the potential risks of receiving them. The limited number of kidney-pancreas, lung, pancreas, and intestinal cases precludes significant conclusions for these organs, though there was no evidence of poor outcomes in these recipients.

Limitations include lack of data on donor infection timeline and estimates of viral load (PCR cycle thresholds), description of donor COVID-19 symptomatology at organ procurement, donor or recipient vaccination or prior COVID-19 infection status, all of which are not tracked in the database. We did not have information that was granular enough to accurately assess transmission of COVID-19 to transplant recipients. Though median follow up times were short for recipients of positive organs, a reflection of how recent these transplants are, there were 141 kidney, 64 liver, and 26 heart transplant recipients with 90 days of follow up. Future analyses of updated versions of the database, which are released quarterly, should address this issue.

### a. Kidneys

COVID-19 severely affected all aspects of renal transplantation. For candidates in the United States, monthly listings dropped 25% and the number of patients in inactive status more than doubled in the first months of the pandemic^12, 13^. In areas with high COVID-19 prevalence, waitlist mortality increased by over 40%^13^. In recipients, COVID-19 infection was found to be devastating: data from the National Health Service (United Kingdom) showed that 26.4% of kidney transplant recipients who tested positive for SARS-CoV-2 died, compared to 10.1% of waitlisted patients^14^. As such, recommendations were made at the time to reserve deceased-donor kidney transplantation for life-saving indications^15^. Advances in COVID-19 diagnostics, therapeutics, and prevention have led to improvement of most of these trends^16^.

Regarding donors, several organizations, including the Association of Organ Procurement Organization, recommended screening deceased donors for SARS-CoV-2 to prevent the inadvertent transplantation of organs from SARS-CoV-2-positive donors. Early in the pandemic, several organizations, including The Association of Organ Procurement Organization, recommended screening deceased donors for SARS-CoV-2 to prevent the inadvertent transplantation of organs from a SARS-CoV-2-positive donor^17, 18^. However, as it became evident to the scientific community that the vast majority of SARS-COV-2 infections are either mild or asymptomatic, and given the significant mismatch in organ supply and demand, accepting organs from deceased donors infected with SARS-CoV-2 was deemed to be viable^19^. Case series of these kidney transplants have since been published, showing good outcomes^20, 21^. Our study, with the biggest number of deceased infected donors and recipients to date, provides reassuring evidence both to patients and the kidney transplant community, that transplanting kidneys recovered from donors who test positive for SARS-CoV-2 within 7 days of organ transplantation is safe.

### b. Livers

Liver transplantation has been severely impacted by the COVID-19 pandemic, particularly in 2020 when the pandemic first emerged in the United States. SRTR data had shown upwards of a 20% reduction in liver transplant listings, 30% reduction in transplants, and 60% increase in waitlist mortality^22^. At that time, balancing the risks and benefits of timely surgery in liver patients who often have an urgent or emergent indication for transplant and no options for bridge therapy versus risk of severe COVID-19 in the post-transplant period was of greatest concern^23^. With the advent of vaccination protocols, antiviral therapies, and improvements in testing and public health policies, transplant trends have been improving as centers are trying to return to baseline rates of liver transplantation^24^.

The issues surrounding donor organ availability are ever-present. They have become more relevant during the pandemic given the high prevalence of SARS-CoV-2 infection in the general population, and thus in the donor pool, and the uncertain safety of transplanting COVID-19-exposed organs into recipients. Guidance from the OPTN and AASLD expert consensus panel have agreed that liver transplantation from donors who recovered from COVID-19 infection and have negative SARS-CoV-2 RNA testing can be safely performed^25, 26^. Reports have emerged with good outcomes in liver transplant recipients whose donors had positive SARS-CoV-2 RNA even up until the time of donation^7, 8, 27, 28, 29^. Using SRTR data, our study provides the best available evidence to date that utilizing organs recovered from donors with SARS-CoV-2 detected within 7 days of liver transplantation is feasible and safe.

### c. Hearts

Similar to other organs, the Covid-19 pandemic has created challenges for the field of heart transplantation (HT) as well. This includes not only the logistical challenges for Organ Procurement Organizations (OPOs) and HT centers but also increased risk of progression and worse outcomes in heart transplant recipients that get infected with COVID-19^30^. Another persistent issue that has not received sufficient attention is the worsening crisis of donor shortage for heart transplantation due to COVID-19. Although there has been an impetus towards finding new avenues for HT donors, like donation after circulatory death (DCD)^31^, use of hepatitis C infected organs, or transplantation of hearts with transient left ventricular dysfunction in order to overcome this shortage, use of donors infected with COVID-19 continues to be an area of uncertainty and many HT-centers may continue to reject such donors largely due to lack of outcome data^32^. Our current analysis using the SRTR data noted equivalent HT outcomes (90-day post HT survival and graft failure) in recipients of hearts from donors with and without COVID-19 infection and adds to the growing body of literature that use of hearts from COVID-19 infected donors is likely safe. While the follow up time for this cohort is short, the current data should provide more reassurance to HT centers when evaluating donors with active COVID-19 infection.

## 7. Conclusions

Overall, our data shows that transplanting kidneys, livers, and hearts recovered from donors recently infected by SARS-CoV-2 is safe. The data also strongly support the notion that donors with recent COVID-19 infection should not be automatically excluded from the donor pool. Prospective studies are needed to confirm our findings and provide insights on optimal post-transplant management of these recipients.

## Data Availability

All data produced in the present work are contained in the manuscript.

## 8. Disclaimer

The data reported here have been supplied by the Hennepin Healthcare Research Institute (HHRI) as the contractor for the Scientific Registry of Transplant Recipients (SRTR). The interpretation and reporting of these data are the responsibility of the authors and in no way should be seen as an official policy of or interpretation by the SRTR or the U.S. Government.

## 9. Disclaimer

These data were presented at the Infectious Diseases Society of New York annual meeting on May 9^th^ 2022.

